# Artificial Intelligence (AI)-based Chatbots in Promoting Health Behavioral Changes: A Systematic Review

**DOI:** 10.1101/2022.07.05.22277263

**Authors:** Abhishek Aggarwal, Cheuk Chi Tam, Dezhi Wu, Xiaoming Li, Shan Qiao

## Abstract

**Background:** Artificial-Intelligence (AI)-based chatbots can offer personalized, engaging, and on-demand health-promotion interventions. This systematic review evaluates the feasibility, efficacy, and intervention characteristics of AI-chatbots in promoting health-behavior change.

**Methods:** A comprehensive search was conducted in seven bibliographic databases (PubMed, IEEE Xplore, ACM Digital Library, PsychoINFO, Web of Science, EMBASE, and JMIR publications) for empirical articles published from 1980 to 2022 that evaluated feasibility and/or efficacy of AI-chatbots for behavioral change. The screening, extraction, and analyses of identified articles followed the PRISMA guidelines.

**Results:** Of the 15 included studies, majority studies (*n*=11) reported high usability, acceptability and engagement, and some evidence on feasibility of AI-chatbots. Selected studies demonstrated high efficacy in promoting healthy lifestyles (*n*=6), smoking cessation (*n*=4), treatment/medication adherence (*n*=2), and reduction in substance misuse (*n*=1). Behavioral change theories and/or expert consultation were used to develop behavioral change strategies of AI-chatbots, including goal setting, monitoring, real-time reinforcement/feedback, and on-demand support. Real-time user-chatbot interaction data, such as user preferences and behavioral performance, were collected on the chatbot platform to identify ways of providing personalized services. The AI-chatbots demonstrated potential for scalability by deployment through accessible devices and platforms (e.g., smartphones and messenger). Participants also reported that AI-chatbots offered a non-judgmental space for communicating sensitive information. However, the reported results need to be interpreted with caution because of moderate to high risk of internal validity, insufficient description of AI-techniques, and limitation for generalizability.

**Conclusion:** AI-chatbots have demonstrated efficacy of health-behavior change interventions among large and diverse population; however, future studies need to adopt robust RCTs to establish definitive conclusions.

## Introduction

Artificial Intelligence (AI)-driven chatbots (AI-chatbots) are conversational agents that mimic human interaction through written, oral, and visual forms of communication channels with a user [1, 2]. With the increased access to technological devices (e.g., smartphones and computers) and internet, AI-chatbots offer the potential to provide accessible, autonomous, and engaging health-related information and services, which can be promising for technology-facilitated interventions. The existing digital therapeutic and telehealth interventions with didactic components, which enable healthcare providers to communicate with patients via digital platforms (e.g., email and online meet), have encountered several challenges including relatively low adherence, unsustainability, and inflexibility [3, 4]. AI-chatbots offer the flexibility of on-demand support, personalized support and content, consistent connectivity (sustainability), and higher interactivity [1,5,6], contributing to addressing the challenges for telehealth interventions.

AI-chatbots demonstrate their potentials through key steps of data processing in health-related conversations: (1) data input, (2) data analysis, and (3) data output. First, AI-chatbots can collect datasets from diverse sources: Electronic Health Records (EHR), unstructured clinical notes, real-time physiological data points (eye-movement tracking, facial recognition, movement tracking, heartbeat), and user interactions [7, 8]. Second, the AI-algorithm uses Machine Learning (ML) and Natural Language Processing (NLP) techniques to identify clinically meaningful patterns and understand user needs [9]. Third, AI-chatbots can mimic real-life human support by offering content or services that can assist users in achieving their health behavior goals [10]. Overall, through acknowledging user needs, demonstrating understanding, and delivering timely services tailored to user preferences (e.g., goal setting, behavioral monitoring, information/knowledge providing), AI-chatbots have the potential to effectively deliver interventions that promote diverse health behaviors (e.g., smoking cessation, physical activity, and medication adherence, etc.). AI-chatbots can also be integrated into embodied functions (e.g, virtual reality), which could provide additional benefits for the health behavior change [11].

In the past decade, evidence regarding the feasibility and efficacy of AI-chatbots in delivering healthcare services has been increasing, while most of these chatbots aim to improve mental health outcomes. Of the extant systematic reviews on AI-chatbots, six articles targeted “ assessing efficacy of AI-chatbots in enhancing mental health outcomes” [1,5,6,9,12,13], two examined ‘feasibility’ of AI-chatbots in health care settings [11, 14], and one described architectures and characteristics of the AI-chatbots used in chronic conditions [15].

Given the merits of AI-chatbots in health promotion, recent literature has paid increasing attention to the use of AI-chatbots for health behavior change. Oh et al. (2021) [2] conducted a systematic review that assessed the efficacy of AI-chatbots for ‘lifestyle modification’ (e.g., physical activity, diet, and weight management). However, the scope and inclusion criteria of this review has several limitations. First, this review did not distinguish AI-driven chatbots and other chabots. For example, the AI-chatbots that performed ‘rule-based’ or ‘constrained’ conversation were included. Second, the selected studies in the existing review targeted a limited set of behaviors including physical activity, diet, and weight management without other types of behaviors. Third, the existing review did not cover all platforms that can deploy AI-chatbots. AI-chatbots that were integrated into virtual reality, augmented reality, embodied agents, and/or therapeutic robots were excluded from the review. Therefore, it is worth doing a systematic review that investigates AI-chatbots integrated into ‘diverse devices’ (robots, smartphones, computers), ‘diverse platforms’ (messenger, virtual/embodied agent, SMS), performed ‘unconstrained’ conversations, and targeted a wide range of behavioral outcomes (smoking cessation, treatment or medication adherence, healthy lifestyle, and related health behavior domains). As such, the current study aimed to conduct a comprehensive systematic review for critically evaluating empirical studies and describing AI-chatbot intervention characteristics, components or functionality, and investigate their feasibility and efficacy in promoting wide range of healthy behaviors.

## Methods

### Data sources and search algorithms

The study protocol of this systematic literature review follows the PRISMA guideline [16] in each step. The comprehensive search was conducted in June 2022 by three authors (C.C. T, S. Q, and A. A) in seven bibliographic databases, including PubMed, IEEE Xplore, ACM Digital Library, PsychINFO, Web of Science, EMBASE, and JMIR publications.

The search was conducted using the combination of various key words from three categories. The first category included keywords related to AI-based chatbot, including “chatbot,” “chatterbot,” “chatter robot,” “artificial intelligence,” “conversational AI,” “conversational agency,” “virtual agent,” “conversational agents,” and “bot”. The second category was related to health behaviors. Keywords in this category included “health promotion,” “health behaviors,” “behavior change,” “substance use,” “alcohol use,” “drinking,” “cigarette use,” “smoking,” “drug abuse,” “drug use disorder,” “risk behaviors,” “lifestyle,” “exercise,” “nutrition behavior,” “sleep,” “adherence,” “body weight,” “physical activity,” “diet,” “risky behaviors,” “healthcare seeking behaviors,” “prescribed medical treatment,” “tobacco use,” and “vaping”. The third category focused on intervention study and included one keyword “intervention”.

Keywords were organized by the following approaches: (1) keywords within one category were lined using the “OR” operator (e.g., “chatbot” OR “conversational AI”); (2) keywords across different categories were connected using the “AND” operator (e.g., “chatbot” AND “health behaviors” AND “intervention”).

### Inclusion and exclusion criteria

The current review selected empirical studies on health behavior interventions applying AI-based chatbot technique according to the following inclusion criteria: (1) intervention research focusing on health behaviors; (2) empirical studies using chatbots; (3) chatbots developed upon existing AI platforms (e.g., IBM Watson Assistant) or AI algorithms, such as machine learning, deep learning, natural language understanding, and natural language processing; (4) studies reporting qualitative or quantitative results on interventions; (5) English articles published from 1980 to 2022 (as of June 2, 2020). Articles were excluded if they were (1) not full-text empirical studies (e.g., conference abstracts or proposals); (2) intervention research with chatbots based on non-AI methods, such as the rule-based approach; (3) studies that did not clarify their AI algorithms; or (4) studies only focusing on mental health but not health behaviors.

As a result, 1961 articles were initially retrieved and screened. A total of 15 articles finally met the inclusion criteria and were selected for the current review (Figure 1). Disagreements in selections were resolved through team discussion.

### Data extraction and quality assessment

Several summary tables were utilized for extracting information from the selected articles, including study characteristics (i.e., author, publication year, study design, participants, age of the sample, sample size, country, and target health behaviors), chatbot-based intervention features (i.e., chatbot types, chatbot components/functionality, settings, existing AI technology, input data sources, platform, theorical foundation, and AI algorithms), and intervention outcomes (i.e., health behavioral outcomes/primary outcomes, feasibility, usability, acceptability, and engagement).

The quality assessment of selected studies was employed in line with the National Institutes of Health (NIH) Quality Assessment Tool for controlled intervention studies [17]. This assessment tool suggests an evaluation of 6 types of risks. Specifically, the risk of reporting outcomes based on ad-hoc analyses was assessed based on prespecified outcomes. The risk of bias in the randomization process was assessed on randomized treatment allocation, concealment of allocation sequence (blinding), and similarity of groups at baselines. The risk of bias due to deviations from the intended interventions was assessed based on concealment of the assigned interventions from the participants, implementors, and evaluators. The risk of outcomes from unintended sources was assessed based on measures to avoid influence of other interventions and fidelity to the intervention protocol. The risk of bias in the measurement of the outcomes was assessed on concealment of assigned intervention from evaluators and validity and reliability of outcome measures. The risk of bias in analysis was assessed on dropout rate, power calculation, and intent-to-treat analysis.

AI techniques specific to AI-chatbot interventions were also appraised using the Consolidated Standards of Reporting Trials-Artificial Intelligence (CONSORT-AI) extension guidance for AI studies [18]. We used a checklist of four domains, including whether the rationale for using AI was specified through the use of AI in context of the clinical pathway; whether the inclusion and exclusion criteria at the level of the input data, and the description of the approaches to handle unavailable input data were specified; whether the input data acquisition processes, and the specifications of human-AI interaction in the collection of input data were described; and whether output of the AI algorithm, and its significance in context of the studies’ outcomes were described. The data extraction and quality assessment were completed by AA and the tables were developed after discussion with SQ.

## Results

### Characteristics of reviewed studies

The characteristics of the reviewed studies are summarized in Table 1. The included journal articles (*n*=15) were published in the following past years: 2 from 2021, 3 from 2020, 6 from 2019, 1 from 2018, 2017, 2013, and 2011 each. Out of 15 studies, 13 reported their geographical location. All 13 studies were distributed across developed countries, with 4 studies from the United States, 2 studies from Australia, and 1 study from each remaining country (Korea, Spain, UK, Japan, France, Swiss, The Netherlands). The sample size in the studies varied from 20 to 99,217 with the median of 116 and the mean of approximately 7200 participants. Six studies had 200+ participants, followed by 4 studies with 100 to 200, 2 studies with 50 to 100, and 3 studies with <50.

**Table 1.** Characteristics of reviewed studies (*n* =15)

| S.no | Study | Study design | Participants | Age of the sample | Sample size | Country | Target health behavior(s)/purposes |
| --- | --- | --- | --- | --- | --- | --- | --- |
| 1 | Piao et al. (2020) [19] | Randomized controlled trial | Office workers | Above 24 years (30-39; 35) | *n* = 106;<br>*n* = 57 (intervention group);<br>*n* = 49 (control group) | Korea | Healthy lifestyle (Physical activity) |
| 2 | Maher et al. (2020) [20] | Pre-post study with no control group | Australian who did not meet Australia physical activity guidelines and not follow a Mediterranean dietary pattern | 45-75 years (56.2) | *n* = 31 | Australia | Healthy lifestyle (Physical activity and healthy diet) |
| 3 | Carrasco-Hernandez et al. (2020) [21] | Randomized controlled trial | Smokers at an outpatient clinic | Average age 49 years | *n* =240<br>*n* = 120 (intervention: chatbot + pharmaceutical treatment)<br>*n* = 120 (control: pharmaceutical treatment) | Spain | Smoking cessation |
| 4 | Stephens et al. (2019) [8] | Pre-post study with no control group | Youths with obesity symptoms at a children’s healthcare system | 9.78-18.54 years (15.20) | *n* = 23 | The United States | Treatment adherence (obesity) |
| 5 | Perski et al. (2019) [22] | Randomized controlled trial | Smokers who purchased the ‘Smoke Free app’ | Equal to and above 18 years | *n* = 6,111<br>*n* = 1061 (intervention: chatbot + “Smoke Free app”)<br>*n* = 5050 (control: “Smoke Free app”) | United Kingdom | Smoking cessation |
| 6 | Masaki et al. (2019) [23] | Pre-post study with no control group | Adult smokers with nicotine dependence | Average age 43.5 years | *n* = 55 | Japan | Smoking cessation |
| 7 | Chaix et al. (2019) [24] | Pre-post study with no control group | Patients with breast cancer | Average age 48 years | *n* = 958 | France | Medication adherence |
| 8 | Calvaresi et al. (2019) [25] | Pre-post study with no control group | Smokers from Facebook communities | NA | *n* = 270 | Swiss | Smoking cessation |
| 9 | Galvão Gomes da Silva et al. (2018) [7] | Post experimental study with no control group (qualitative study) | Volunteers from School of Psychology’s pool | Above 18 years (18-25; 23) | *n* = 20 | N/A | Healthy lifestyle (physical activity) |
| 10 | Stein & Brooks (2017) [10] | Pre-post study with no control group | Overweight and obese adults (body mass index ≥ 25) | Average age 47 years | *n* = 70 | The United States | Healthy lifestyle (weight loss, healthy dietary, physical activity, healthy sleep duration) |
| 11 | Crutzen et al. (2011) [26] | Pre-post study with no control group | Adolescents interested in the intervention | Average age 15 years | *n* = 920 | The Netherlands | Healthy lifestyle |
| 12 | Brar Prayaga et al. (2019) [27] | Cross-sectional study (post study) | Medicare recipients | Average age 71 years | *n* = 99,217 | The United States | Medication adherence |
| 13 | Prochaska et al. (2021) [28] | Pre-post study with no control group | American adults screened positive for substance misuse | Above 18 years (18-65; 36.8) | $n = 101$ | The United States | Reducing problematic substance use |
| 14 | To et al., (2021) [29] | Quasi-experimental design without a control group | Inactive individuals (<20 min/day of moderate-to-vigorous physical activity), | Average age 49.1 years | $n = 116$ | Australia | Healthy lifestyle (Physical activity) |
| 15 | Bickmore et al. (2013) [30] | Randomized controlled trial (4-arm) | Individuals in pre-contemplation or Contemplation Stages of Change with respect to moderate-or-greater intensity physical activity or consumption of fruits and vegetables. | 21 - 69 years (33) | $n = 122$ | NR | Healthy lifestyle (Physical activity and healthy diet) |

Out of 14 studies that reported mean age of participants, most studies had adult participants aged 18-30 years (*n*=2), 30-40 years (*n*=3), 40-50 years (*n*=5), 50-60 years (*n*=1), and 60+ years (*n*=1), with only 2 studies having participants with <18 years of age. The selected studies had participants with diverse pre-existing conditions: individuals with lower physical exercise and healthy diet levels (*n*=4), smokers (*n*=4), obese patients (*n*=2), breast cancer patients (*n*=1), substance misuse (*n*=1), general population (*n*=2), and Medicare recipients (*n*=1). The target health behaviors of studies included promotion of healthy lifestyle (physical exercise, diet, *n*=5), smoking cessation (*n*=4), treatment or medication adherence (*n*=3), and reducing problematic substance use (*n*=1). Only 4 studies used randomized control trials (RCTs), and majority of studies (*n*=9) adopted a quasi-experimental design (i.e., pre- and post- tests) with no control group, followed by 1 study with a cross-sectional design and 1 study with a post experimental research method.

### Intervention study quality assessment

The results of quality assessment were presented in Table 2. *The risk of reporting outcomes* was low as all the studies prespecified their outcomes and hypothesis. *The risk of bias in the randomization process* was low. All 4 RCTs adopted appropriate randomized treatment allocation and reported concealment of allocation sequence from participants, and 3 RCTs established similarity of groups at the baseline. The non-RCT studies (*n*=11) were not applicable for the assessment of the randomization process.

**Table 2.**
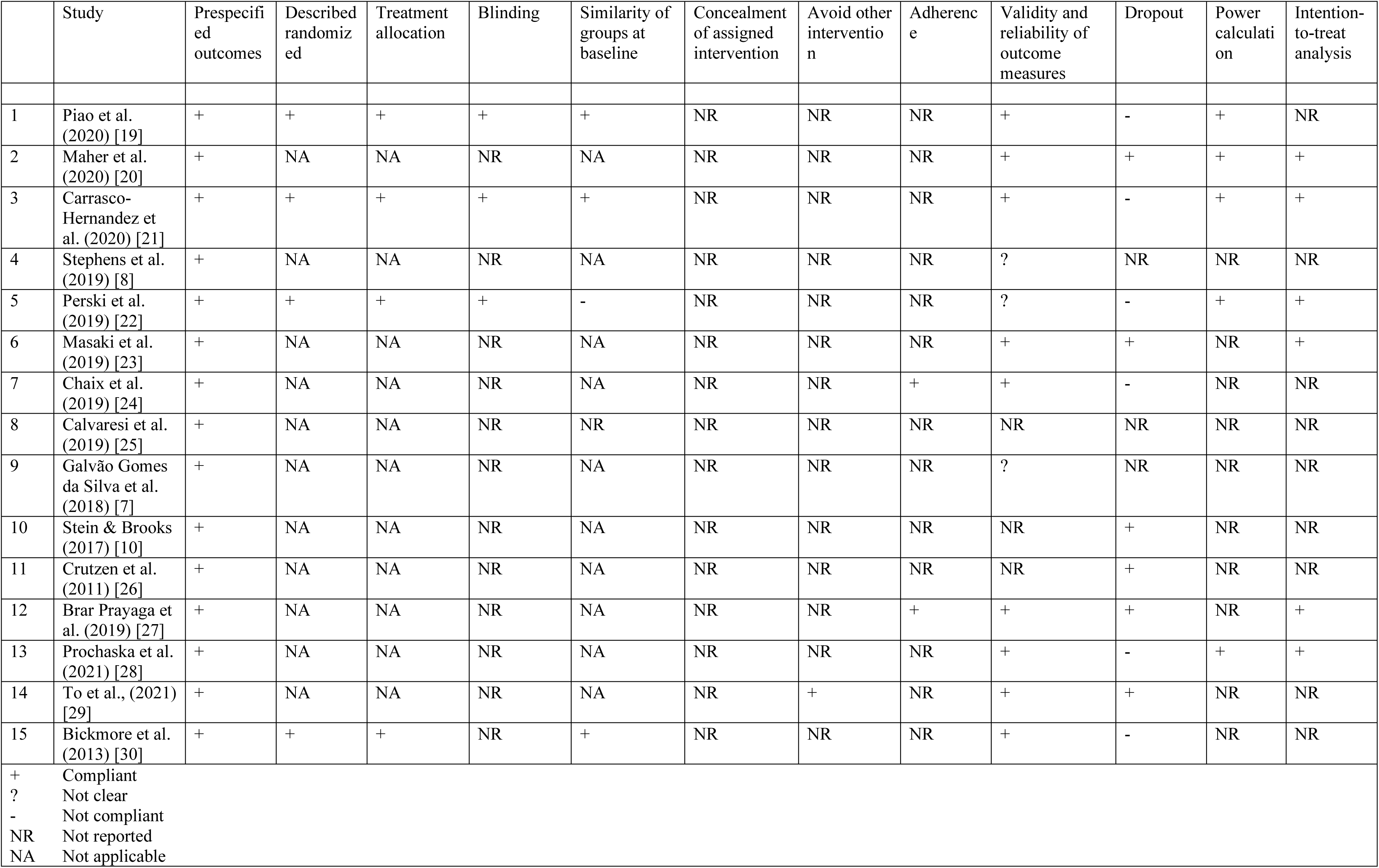
Methodology assessment based on the NIH quality assessment tool for controlled intervention studies

*Risk of bias of deviations from the intended interventions* was considered low to moderate. Although none of the studies (*n*=15) reported concealment of the assigned interventions from facilitators, evaluators, and participants, it was mainly because concealment from the persons providing and receiving behavioral, lifestyle, or surgical interventions is difficult [17]. *Risk of outcomes from unintended sources* was high. First, none of the studies reported any explicit measures to avoid the influence of other interventions on the outcomes or the existing intervention. In case of RCTs (*n*=4), this bias is minimized due to the experimental setting of the interventions; however, for non RCTs (*n*=11), there was high risk of bias due to the potential effect of confounding variables. Second, most of the studies (*n*=13) did not report whether participants adhered to the intervention protocols.

*Risk of bias in the measurement of the outcomes* was moderate. First, none of the studies reported whether the assigned intervention was concealed from the evaluators. Second, nine studies reported the reliability and validity of the outcome measures. In the remaining studies, the reliability and validity of outcome measures was not clear (*n*=3) or not reported (*n*=3). *Risk of bias in analysis* was moderate to high. First, studies with a ≥15% differential dropout rate between groups, and ≥20% dropout rate for intervention or control group were considered to have high dropout rate [17]. Only 5 studies had a lower dropout rate than the cutoff limits, 3 studies did not report the dropout rate, and 7 studies had a higher dropout rate than the cutoff limits. Second, only 5 studies reported the use of power calculation to estimate a sample size that can detect a significant difference in primary outcomes. Third, only 6 studies adopted intent-to-treat analysis.

### AI quality assessment

The AI component of the chatbots was evaluated to demonstrate AI’s impact on health outcomes (Table 3). *Rationale for using AI* was prespecified in all studies (*n*=15). *Characteristics and handling of the input data for AI* was described in no study except one. *Input data acquisition processes for AI* was mentioned in all studies except two. *Specifications of the human-AI interaction* was reported in the collection of input data in majority of the studies (*n*=9). *AI algorithms’ output and its significance in context of the studies’ outcomes* was described in all studies except two. Conclusively, there was sufficient description for all factors except for input data characteristics and handling of unavailable input data.

**Table 3.**
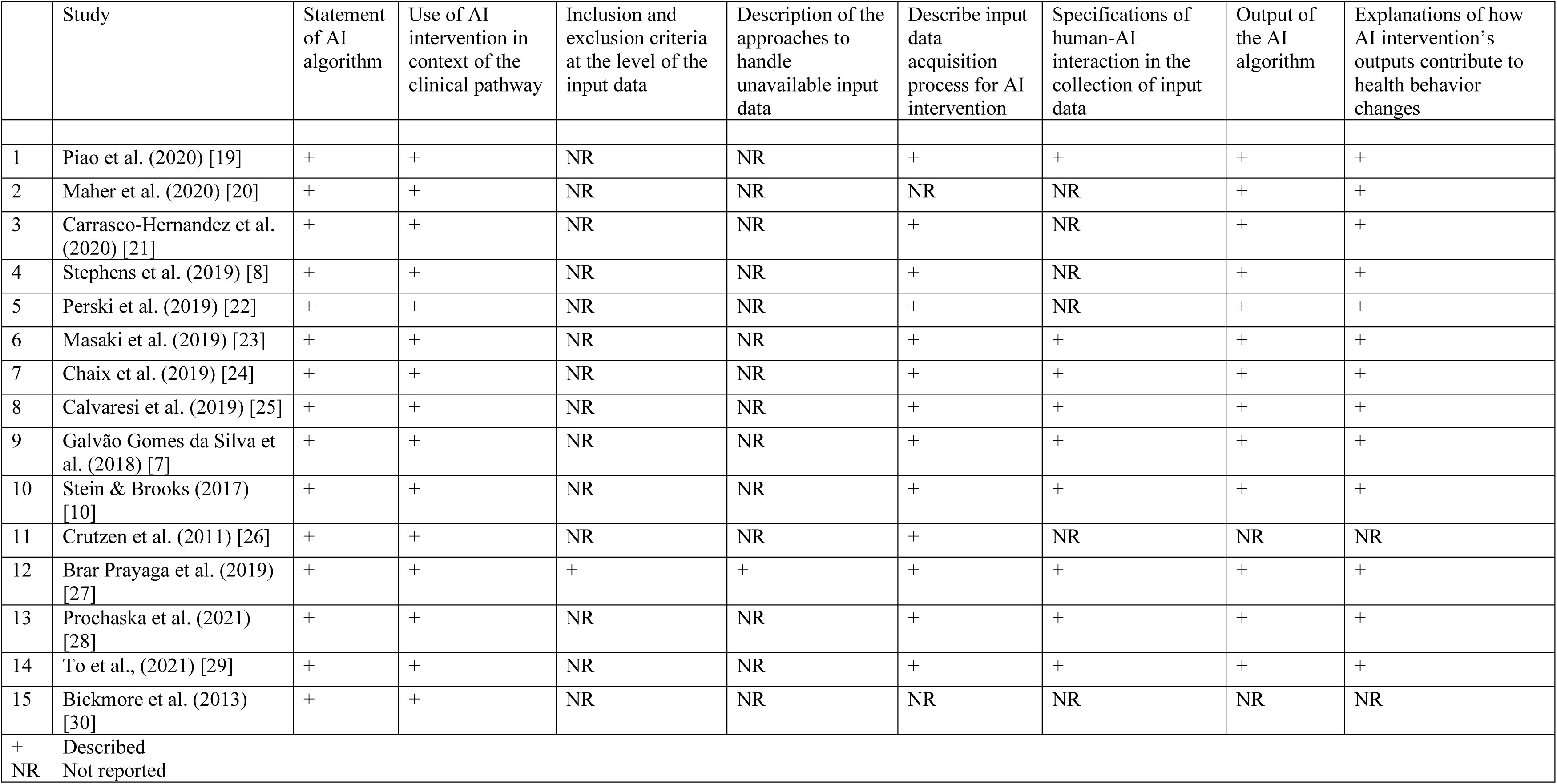
Quality assessment on Chatbot interventions based on CONSORT guidance – AI extension

### Outcomes of Reviewed Studies

#### Feasibility

The outcomes of the selected studies are reported in Table 4. Out of 15 studies, 6 studies reported feasibility of AI-chatbots in terms of: (1) safety, and (2) messages exchanged with the chatbot. First, Maher et al. (2020) [19] reported feasibility in terms of safety (i.e., no adverse events were reported). Second, the remaining three studies reported feasibility as the total and *Mean (M*) number of messages exchanged with the chatbot per month. The maximum number of message exchanges per month were 132970 (*M* = 139) [20], followed by 4123 (*M* = 90) [8], 42217 (*M* = 11.3) [21], 600 messaged sent (*SD* = 556.5) [22], and average 6.7 (*SD* = 7.0) messages per week [23]. Stephens et al. (2019) [8] also reported that the chatbot conversation enabled participants to progress towards their goals 81% of the time. Overall, there was very less evidence on the safety of chatbots, and some evidence on the feasibility of chatbots in terms of total and mean number of messages exchanged.

**Table 4.** Outcomes of reviewed articles

| Study | Feasibility | Usability | Acceptability and Engagement | Health behavior outcomes (primary outcome) |
| --- | --- | --- | --- | --- |
| Piao et al. (2020) [19] | | | | (1) Habit strength (Self-Report Habit Index (SRHI)) (intervention vs. control)<br>Results:<br>- Significant between-group differences in SRHI when controlled for intrinsic award via app ($p=.008$)<br>- Non-significant between-group differences in SRHI, apart from physical activity ($p=.045$), when not controlled for intrinsic rewards via app ($p=.21$)<br>Analytical methods and results: Repeated measures analysis of variance (ANOVA) |
| Maher et al. (2020) [20] | (1) *Safety*<br>Results: No adverse events were reported. | (1) *Ease of Use*<br>Results: Participants had minimal smartphone skills<br>(2) *Content usability*<br>Results: Difficulty in consuming the recommended number of food group servings | (1) *Retention rate*<br>Results: 5 withdrawals (83.9% retention rate)<br>(2) *Duration of engagement*<br>a) 64% check-ins completed on average by participants<br>b) Decrease in check-ins by 20% (mid program) followed by an increase in final weeks | (1) Physical activities (total minutes of weekly physical activity; Active Australia Survey) (week 12 vs baseline)<br>Results: *Mean* improvement 109.8 minutes [95% *CI* (1.9, 217.7)] [$F(2, 29) = 6.45, p = .005$]<br>(2) Adherence to healthy (Mediterranean) diet (Australian Mediterranean diet adherence tool) (week 12 vs baseline)<br>Results: *Mean* improvement by 5.7 points [95% *CI* (4.2, 7.3)] [$F(2,29) = 44.45, p < .001$]<br>(3) Weight (Kg) (week 12 vs baseline)<br>Results: Decrease in the average weight by 1.3 Kgs [95% *CI* (−0.1, −2.5)] [$F(2, 29) = 5.41, p = .01$]<br>(4) Waist circumference (cm) (week 12 vs baseline)<br>Results: Decrease in the average waist circumference by 2.1 cm [95% *CI* (−3.5, −0.7)] [$F(2, 29) = 7.13, p = .003$]<br>(5) Systolic and diastolic blood pressure (Omron Healthcare)<br>Results: Mean improvement 0.2 [$F(2,29) = 0.11, p = .90$], and 1.0 [$F(2,29) = 0.64, p = .54$], respectively.<br>Analytical methods: Repeated measures ANOVA |
| Carrasco-Hernandez et al. (2020) [21] | | (1) *Content usability* (Perceived quality questionnaire, range: 1 to 5)<br>Results: averagely high scores: E.g., “I felt in control of modifying my interest profile” ($Mean = 3.63, SD = 0.66$); “The messages recommended to me were diverse” ($Mean = 4.47, SD = 0.51$) | (1) *Duration of engagement* (the ratio of rated messages divided by the total number of message)<br>Results: Engagement was the highest at first month and reduced gradually, becoming lowest at 12 months. A significant difference was found in system engagement at 6 months ($p=.04$) but not in all subsequent months. | (1) Smoking abstinence rate (exhaled carbon monoxide and urine cotinine tests)<br>Results: *OR* = 2.15 [95% *CI* (1.13, 4.08), $p = .02$] (intervention vs. control; 12 months)<br>Analytical method: Multinomial model<br>(2) Health-related quality of life (Short-Form Health Survey and EuroQo1)<br>Results: *OR* = 0.17 [95% *CI* (-6.36, 6.70)] (intervention vs. control; at 6 months); *OR* = 3.01 [95% *CI* (-3.14, 9.16)] (at 12 months) |
|  |  |  |  | (3) Healthy lifestyle (body mass index)<br>Results: $OR = -0.09$ [95% $CI (-0.77, 0.60)$, $p = .80$] (intervention vs. control; at 6 months); $OR = 0.25$ [95% $CI (-0.53, 1.03)$, $p = .52$] (at 12 months)<br>(4) Physical activity (International Physical Activity Questionnaire)<br>Results: $p = .47$ (at 6 months); $p = .73$ (at 12 months) (intervention vs. control)<br>Analytical methods:<br>Multinomial model and chi-square test |
| Stephens et al. (2019) [8] | (1) *Messages exchanged*<br>Results:<br>The total number of conversations with chatbot per month<br>Results: 4123 ($Mean = 90$ conversations per person)<br>(2) *Target goal progress* (percent times progressed toward the target goals)<br>Results: 81% of the time | (1) *Content usability* (self-reported endorsement of helpfulness during the conversation):<br>Results: 96% of the time<br>(2) *Proportion of user-initiated conversation* (ratio of Chatbot-initiated vs. patient-initiated conversations)<br>Results: 26.4% were patient-initiated<br>(2) *Outside office support*<br>Results: Support provide outside office (sustainability)<br>Results: 17.8% of total support (55 hours and 45 minutes) | (1) *Duration of engagement* (duration of conversations)<br>Results: average duration of conversation ($Mean = 12.5$ minutes, $SD = 15.62$) |  |
| Perski et al. (2019) [22] |  |  | (1) *Duration of engagement* (automatically recorded number of logins) (intervention vs. control)<br>Results: $Median = 16$, $IQR = 65.5$ (intervention) vs. $Median = 5$, $IQR = 22$ was associated with 107% increase in engagement ($p < .001$)<br>Analytical methods: Negative binomial regression analyses | (1) Smoking abstinence (continuous abstinence)<br>Results: Intervention group had 2.44 times greater odds of being abstinent. $AOR = 1.36$ [96% $CI (1.16, 1.61)$, $p < .001$]<br>Analytical methods: logistic regression |
| Masaki et al. (2019) [23] |  | (1) *Content usability*<br>Results:<br>a) Number of times "Like!" tapped to the advice provided by the chatbot (from week 0 to 12)<br>Results: $Mean = 26.5$ times ($SD = 63.8$)<br>b) Number of times “Call” tapped to for AI nurse in context of smoking impulses or side effects (from weeks 0 to 12)<br>Results: $Mean = 1.7$ times ($SD = 2.4$) | (1) *Duration of engagement*<br>Results:<br>a) Number of days of diary entries (from weeks 0 to 12)<br>Results: $Mean = 56.1$ ($SD = 31.3$)<br>b) Number of educational videos viewed from start to finish<br>Results: $Mean = 12.6$ ($SD = 6.8$) | (1) Smoking abstinence (continuous abstinence rate)<br>Results: 76% [95% $CI (65, 88)$] (12 weeks), 64% [95% $CI (51, 76)$] (24 weeks), 58% [95% $CI (46, 71)$] (52 weeks)<br>(2) Cigarette withdrawal (Mood and Physical Symptoms scale)<br>Results: Differences = -6.4 ($SD = 5.8$) (12 weeks – baseline)<br>(3) Tobacco craving (12-item French version of the Tobacco Craving Questionnaire)<br>Results: Differences = -0.6 ($SD = 1.5$) (12 weeks – baseline) |
|  |  |  |  | (4) Social nicotine dependence (Kano Test for Social Nicotine Dependence)<br>Results: Differences = -6.7 (*SD* = 5.2) (12 weeks – baseline) |
| Chaix et al. (2019) [24] | (1) *Messages exchanged*<br>The total number of conversations with chatbot per month<br>Results: 132970 (*Mean* = 139 conversations per person) | (1) *Content usability*<br>Results:<br>a) Rate of participants who reported satisfaction with the use of chatbot<br>Results: 93.95% (900/958)<br>b) Rate of participants who reported that chatbot was supportive or helpful to track their treatment effectively<br>Results: 88% (943/958)<br>(2) *Non-judgmental safe space*<br>Results: Share personal and intimate information such as sexuality. | (1) *Retention rate* (users who send at least one message per month)<br>Results: 72% (2nd month) reduced to 31% (8<sup>th</sup> month) | (1) Medication adherence rate (The average compliance of patients using the prescription reminder feature)<br>Results: More than 20% improvement in 4 weeks (*p* = .04)<br>Analytical methods: *t*-test |
| Calvaresi et al. (2019) [25] |  |  | (1) *Duration of engagement* (rate of participants operating the app on social network)<br>Results: 74% (200/270) users engaged actively | (1) Rate of participants who succeeded in the smoking cessation goal, three months after the last cigarette<br>Results: 28.9% (78/270) participants succeeded in the smoking cessation goal<br>(2) Rate of the improvement on obtained cessations compared to previous campaign without a chatbot<br>Results: 10% |
| Galvão Gomes da Silva et al. (2018) [7] |  | (1) *Easy of use*<br>Results: Easy to tap on the robot’s head to continue the conversation.<br>(2) *Content usability*<br>Results: Clear and easy to understand interview script, but with some ambiguity.<br>(3) *Non-judgmental safe space*<br>Results: Chatbot offered space and time to talk |  | Qualitative data<br>(1) Interview evaluation<br>- Connection: Tension or awkwardness, followed by comfortable experience.<br>(2) Motivation<br>- Enhanced immediate motivation<br>(3) Engagement of physical activity after the program<br>- Mixed reports<br>(4) Positive features<br>- Flexible and increased self-awareness and social skills. |
| Stein & Brooks (2017) [10] |  | (1) *Content usability*<br>Results:<br>a) In-app user trust survey<br>Results: 100% response rate and positive results<br>b) Average satisfaction score<br>Results: 87 out of 100<br>c) Average net promoter score (promoters (9-10) – detractors (0-6))<br>Results: 47 | (1) *Duration of engagement* (time in weeks between first and final use of the app)<br>Results: The average duration of app use was 15 weeks (*SD* 1.0), and users averaged 103 sessions each. | (1) Weight loss (percent weight change)<br>Results: Weight loss (standard error of the mean) was 2.38% (0.69%) of baseline weight in 75.7% (53/70) of users.<br>(2) Meal quality (if they contained healthy food and no unhealthy food)<br>Results: Percentage of healthy meals increased by 31%, and the percentage of unhealthy meals decreased by 54%. |
|  |  | d) Average scores for disappointment if app not offered<br>Results: 6.73 |  |  |
| Crutzen et al. (2011) [26] | (1) *Messages exchanged*<br>Results: The total number of conversations with chatbot per month<br>Results: 42217 (*Mean*= 11.3 conversations per person) | (1) *Ease of use* (visual analog scale)<br>Results: Low (N = 852, M =47.8, *SD* =31.4)<br>(2) *Content usability*<br>Results:<br>a) Reliability of the information provided by chatbot<br>Results: High (N = 852, M =56.4, *SD* =51.5)<br>b) Chatbot was considered easier to use as compared with information lines<br>c) Chatbot was considered to provide better quality and concise of information in comparison with search engines.<br>d) The quantity of information of the chatbot was considered less in comparison with both information lines and search engines<br>(3) *Non-judgmental safe space*<br>Results: Chatbot was used for questions regarding sex, drugs, and alcohol, and was considered more anonymous and faster than information lines | (1) *Duration of engagement*<br>Results:<br>a) The average duration of chatbot use per person = 45 min<br>b) The average duration of a conversation per person<br>Results: *Mean* = 3 minutes 57 seconds |  |
| Brar Prayaga et al. (2019) [27] | | | | (1) Refill requests from participants (rate = requests/total reminders)<br>Results: 17.40% (47,552/273,356)<br>(1) Language (Spanish vs. English)<br>Results:<br>- Spanish-speakers had significantly lower refill request rates than English-speaking patients (10.69% vs. 18.83%) ($\chi^2 = 1829.2$; $p < .001$)<br>- Among these who engaged with the reminder, Spanish-speaking patients had higher refill request rates than English-speaking patients (57.31% vs. 49.22%, $\chi^2 = 212.5$, $p < .001$)<br>(2) Age (< 45 years vs. > 75 years)<br>Results: Younger patients had higher refill request rates than older patients (47.81% vs. 29.84%, $\chi^2 = 1460.3$, $p < .001$)<br>Analytic methods: chi-square test ($\chi^2$) |
| Prochaska et al. (2021) [28] | (1) *Messages exchanged*<br>Results: 600.7 (SD 556.5) sent messages. | (1) *Content usability*<br>Results:<br>(a) Fewer felt Woebot met most or all of their needs (22/51, 43%). | (1) *Duration of engagement*<br>Results:<br>(a) Participants' W-SUDs use averaged 15.7 (SD 14.2) days, 12.1 (SD 8.3) modules.<br>(b) About 94% (562/598) of all completed psychoeducational lessons were rated positively.<br>(2) *Acceptability*<br>(a) Most participants reported receiving the service they desired (41/51, 80%) and would recommend W-SUDs to a friend (39/51, 76%). | (1) Significant increase in confidence to resist urges to use substances (mean score change +16.9, SD 21.4; $p<.001$)<br>(2) Significant decrease in substance use occasions (mean change -9.3, SD 14.1; $p<.001$), Alcohol Use Disorders Identification Test-Concise (mean change -1.3, SD 2.6; $p<.001$), 10-item Drug Abuse Screening Test (mean change -1.2, SD 2.0; $p<.001$), Patient Health Questionnaire-8 item (mean change 2.1, SD 5.2; $p=.005$), Generalized Anxiety Disorder-7 (mean change 2.3, SD 4.7; $p=.001$), and cravings scale (68.6% vs 47.1% moderate to extreme; $p=.01$). |
| To et al., (2021) [29] | (1) *Message frequency*<br>(a) On average, 6.7 (SD 7.0) messages/week were sent to the chatbot.<br>(b) About half of the participants (58/113) sent messages to the chatbot at least once a day. | (1) *Content usability:*<br>Results:<br>(a) Most of the participants scored overall the usability of the chatbot (101/113, 89.4%) as at least "OK" (System Usability Scale).<br>(b) Average usability score for the chatbot was (49/113, 43.4%), including increase in confidence (52/113, 46%), overcoming barriers (60/113, 53.1%), increased support (59/113, 52.2%), planning (63/113, 55.7%), staying motivated (47/113, 41.6%), and becoming more active (60/113, 53.1%). | (1) *Duration of engagement*<br>(a) Participants spent 5.1 minutes with the chatbot per day.<br>(b) 71/113, 62.8% participants always read the chatbot messages.<br>(2) *Satisfaction*<br>(a) About one-quarter liked very much the messages that the chatbot sent out (26/113, 23%).<br>(b) Only 43.4% (49/113) thought that the chatbot understood their messages most of the time.<br>(c) About one-third would continue to use the chatbot in the future (40/113, 35.4%).<br>(3) *Technical issues*<br>Most participants experienced technical issues (93/113, 82.3%) and stopped receiving the chatbot messages at any time during the study (95/113, 84.1%). | At follow-up, results:<br>(1) Participants recorded more steps (increase of 627, $p<.01$) and,<br>(2) More total physical activity (increase of 154.2 min/week; 3.58 times higher at follow-up, $p<.001$).<br>(3) The decrease in BMI was not significant (-0.13 *CI* -0.37 to 0.11)<br>(4) Participants were also more likely to meet the physical activity guidelines (*OR* 6.37, 95% *CI* 3.31-12.27) at follow-up. |
| Bickmore et al. (2013) [30] | - | (1) *Ease of use*<br>(a) Participants reported relative ease of using the chatbot *Mean* = 4.80 (1.97) on a scale of 1 to 7. | (1) *Satisfaction*<br>(a) Participants were above average satisfied with agent *Mean* = 4.30 (1.84) on a scale of 1 to 7.<br>(b) Participants reported below average desire to continue with agent *Mean* =3.75 (2.18) on a scale of 1 to 7. | (1) No significant differences among conditions in International Physical Activity Questionnaire (IPAQ) ($F(3,107)=1.07$, $p=0.367$).<br>(2) A significant difference between conditions with the DIET intervention doing the best ($F(3,103)=4.52$, $p=0.005$).<br>(3) No significant differences among conditions for weight ($F(3,105)=1.09$, $p=0.374$). |

#### Usability

Out of 15 studies, 11 studies reported usability of AI chatbots in terms of: (1) ease of using the chatbot, (2) outside-office support, (3) proportion of user-initiated conversation, (4) usability of the content, and (5) non-judgmental safe space. First, 4 studies reported the ‘perceived’ ease of using chatbots by the participants. Maher et al. (2020) [19] reported that the participants did not have sufficient smartphone skills to operate the chatbot. Likewise, Crutzen et al. (2011) [21] reported that the ease of using chatbot on the visual scale was low (*n* = 852, *M* = 47.8, *SD* = 31.4); however, the chatbot was still considered easier to use as compared to information lines. Galvão Gomes da Silva et al.’s (2018) [7] reported that their social robot, NAO, was considered easy to use as the participants felt comfortable tapping on the robot’s head to move to the next question. Bickmore et al. (2013) [24] reported above average ease of use (*Mean* = 4.80, *SD* = 1.97) on a scale of 1 to 7. Overall, the ease of using chatbots was dependent on the platform, user interface, and the cultural sensitivity in chatbot’s design. Second, only Stephens et al. (2019) [8] reported that use of chatbot was not limited to the office, 17.8% of total support (55 hours and 45 minutes) was provided outside the office, ensuring a consistent and sustained connection with participants. Third, Stephens et al. (2019) [8] also reported that 26.4% of the conversations were user-initiated.

Fourth, majority of the studies (*n*=10) demonstrated the usability of the content shared by the chatbot through self-report measures and the number of times chatbot services were used. For total duration of 12 weeks, Masaki et al. (2019) [25] reported the number of “likes” to the advice provided by the chatbot (*M* = 26.5 times, *SD* = 63.8), and the number of “calls” to the AI nurse to seek assistance for smoking impulses or side effects (*M* = 1.7 times, *SD* = 2.4). Crutzen et al. (2011) [21] reported that Bzz’s content was considered more reliable (*n*=852, *M* =56.4, *SD* =51.5), concise, and of higher quality as compared to the content of search engines and information lines; however, the quantity of information by the chatbot was considered lesser. Galvão Gomes da Silva et al.’s (2018) [7] reported that NAO’s interview script was considered clear and easy to understand, but with scope of improvement to remove ambiguity. Carrasco-Hernandez et al. (2020) [26] reported on average high scores on different factors in the perceived quality questionnaire (Range: 1 to 5) – personalized messaged (*M* = 3.63, *SD* = .66), diverse information (*M* = 4.47, *SD* = .51), etc.

Through the in-app trust survey with a 100% response rate, Stein & Brooks (2017) [10] reported high satisfaction score (87/100), high net promoter score (8.3) [subtracting the promoters (9-10) from detractors (0-6)], and high disappointment level if the chatbot was not offered (6.73). Stephens et al. (2019) reported that 96% of the times the chatbot content was endorsed as helpful by the participants. Chaix et al. (2019) [20] reported high user satisfaction (93.95% users, total *n* = 958) and helpfulness of the chatbot to track their treatment effectively (88% users, total *n* = 958). Maher et al. (2020) [19] reported that participants experienced difficulty in consuming the recommended number of food group servings. Prochaska et al., (2021) [22] reported that fewer participants felt Woebot met most or all of their needs (22/51, 43%). To et al., (2021) reported that most of the participants scored overall the usability of the chatbot (101/113, 89.4%) as at least “OK” (System Usability Scale), with the chatbot assisting in increasing confidence (46%), overcoming barriers (53.1%), increased support (52.2%), planning (55.7%), staying motivated (41.6%), and becoming more active (53.1%).

Fifth, 3 studies reported that the AI-platforms offered a non-judgement safe space for users to share detailed and sensitive information. Galvão Gomes da Silva et al.’s (2018) [7] NAO was considered non-judgmental social robot that provided participants with personal space and time to think and respond uninterruptedly. Though initially participants felt awkward while interacting with NAO, gradually, they felt relaxed or comfortable and found the interaction interesting, surreal, or unusual. Chaix et al. (2019) [20] reported that Vik offered a platform for the users to share personal and intimate information such as sexuality which they could not share with the doctor directly. Likewise, Crutzen et al. (2011) [21] reported that 48% of the adolescents preferred the chatbot for questions regarding sex, drugs, and alcohol over information lines and search engines, and the chatbot was considered more anonymous and faster than information lines. Conclusively, the chatbot content was considered highly useful in achieving behavioral goals.

#### Acceptability and Engagement

Out of 15 studies, 12 studies reported acceptability and engagement of AI chatbots in terms of: (1) retention rate, and (2) duration of engagement, (3) satisfaction, and (4) technical issues. First, 2 studies reported the retention rate. Chaix et al. (2019) [20] reported a gradual decrease in the retention rate (users who send at least one message per month over 8 months from 72% (2^nd^ month) to 31% (8^th^ month), whereas, Maher et al. (2020) [19] reported 83.9% retention rate (users who completed the study). Second, 8 studies reported duration of engagement through diverse factors. Carrasco-Hernandez et al. (2020) [27] calculated engagement as the proportion of rated messages. Engagement was the highest at first month and reduced gradually, becoming lowest at 12 months. A significant difference was found in system engagement between groups at 6 months (*p* = .04), but not in all subsequent months. Maher et al. (2020) [19] reported a decrease in check-ins by 20% mid program, followed by an increase to 70% in the final week, with average number of logins completed by participants at 64%. Likewise, Calvaresi et al. (2019) [28] reported the rate of active users 74% (*n*=270).

Perski et al. (2019) [29] reported a significant difference in the engagement levels (automatically recorded number of logins) between group [*Median* = 16, *IQR* = 65.5 (intervention group) vs. *Median* = 5, *IQR* = 22 (control group)] that was associated with a 101% higher engagement in the group that used chatbot (intervention group) (*p* < .001). Stephens et al. (2019) [8] and Crutzen et al. (2011) [21] reported the average duration of conversation as 12.5 minutes (*SD* = 15.62) and 4 minutes (average duration of chatbot use as 45 minutes), respectively. Masaki et al. (2019)’s [25] reported the number of days of diary entries (*M* = 56.1, *SD* = 31.3), and the number of educational videos viewed from start to finish (*M* = 12.6, *SD* = 6.8). Stein & Brooks (2017) [10] reported that the average duration of app use was 15 weeks (*SD* = 1.0), averaging 103 sessions per user. Prochaska et al. (2021) [22] reported that participants’ Woebot usage averaged 15.7 (*SD* 14.2) days, 12.1 (*SD* 8.3) modules, and about 94% (562/598) of all completed psychoeducational lessons were rated positively. To et al., (2021) [23] reported that participants spent 5.1 minutes with the chatbot per day and 62.8% (71/113) participants always read the chatbot messages.

Second, To et al., (2021) [23] reported that about one-quarter of the participants liked very much the messages sent by the chatbot (26/113, 23%); but, only 43.4% (49/113) thought that the chatbot understood their messages most of the time. About one-third would continue to use the chatbot in the future (40/113, 35.4%). Bickmore et al. (2013) [24] reported that participants were above average satisfied with agent (*M* = 4.30, *SD* = 1.84 on a scale of 1 to 7), reported below average desire to continue with agent in future (*M* =3.75, *SD* = 2.18) on a scale of 1 to 7. Third, To et al., (2021) [23] reported that most participants experienced technical issues (93/113, 82.3%) and stopped receiving the chatbot messages at any time during the study (95/113, 84.1%). Conclusively, though chatbots contributed to user engagement significantly, there was an indication of decrease in the engagement rate with time.

#### Efficacy

##### Healthy lifestyle (physical exercise and diet)

Out of the 15 studies, 7 studies targeted healthy lifestyles, and 6 studies assessed the efficacy of AI chatbots in promoting healthy lifestyles through: (1) physical activity levels, (2) healthy diet, (3) motivation, (4) blood pressure, and (5) BMI. First, all 6 studies except one reported an increase in the physical activity. Stein & Brooks (2017) [10] reported that increase in physical activity led to an average weight loss of 2.38% in 75.7% (*n*=70) of the users. Maher et al. (2020) [19] reported an increase in the physical activity by 109.8 minutes [95% *CI* (1.9, 217.7)] [*F*(2, 29) = 6.45, *p* = .005], and a decrease in the average weight and waist circumference by 1.3 Kgs [95% *CI* (–0.1, –2.5)] [*F*(2, 29) = 5.41, *p* = .01] and 2.1 cm [95% *CI* (–3.5, –0.7)] [*F*(2, 29) = 7.13, *p* = .003]. Piao et al. (2020) [30] reported significant between-group differences in Self-Report Habit Index (SRHI) when controlled for intrinsic reward via chatbot enables app (*p* = .008). Galvão Gomes da Silva et al.’s (2018) [7] qualitative analysis reported mixed results on achieving goals on physical activity. To et al., (2021) [23] reported that participants recorded more steps (increase of 627, *p*<.01) and more total physical activity (increase of 154.2 min/week; 3.58 times higher at follow-up, *p*<.001). Moreover, participants were also more likely to meet the physical activity guidelines (*OR* 6.37, 95% *CI* 3.31-12.27) at follow-up. However, only Bickmore et al. (2013) [24] reported no significant differences among conditions in International Physical Activity Questionnaire (IPAQ) (*F*(3,107)=1.07, *p*=0.367).

Second, 3 studies reported an improvement in diet. Stein & Brooks (2017) [10] reported that percentage of healthy meals increased by 31% and the percentage of unhealthy meals decreased by 54%. Maher et al. (2020) [19] reported an increase in the mean of Mediterranean diet (healthy meal) scores by 5.7 points [95% *CI* (4.2, 7.3)] [*F*(2,29) = 44.45, *p* < .001]. Bickmore et al. (2013) [24] reported that a significant difference between conditions with the DIET intervention doing the best (*F*(3,103)=4.52, *p*=0.005), but no significant differences among conditions for weight (*F*(3,105)=1.09, *p*=0.374). Third, Galvão Gomes da Silva et al.’s (2018) [7] reported enhanced immediate motivation towards activities such as planning, goal setting, meeting friends and families, and increasing will power through mindfulness techniques. Participants also reported that they felt more self-aware, and open to share their goals with others. Fourth, Maher et al. (2020) [19] assessed blood pressure level post intervention as a secondary outcome; however, the mean improvement in systolic blood pressure (0.2) [*F*(2,29) = 0.11, *p* = .90] and diastolic blood pressure (1.0) [*F*(2,29) = 0.64, *p* = .54] was not significant. Fifth, only To et al., (2021) [23] reported that the decrease in BMI was not significant (-0.13 *CI* -0.37 to 0.11). Conclusively, there were significant differences in the primary outcomes of interest (physical activity levels, and healthy diet) in all studies aimed at improving healthy lifestyles.

##### Smoking Cessation

Out of the 15 studies, 4 studies assessed the efficacy of AI chatbots in smoking cessation. Perski et al. (2019) [29] reported that the intervention group had 2.44 times greater odds of abstinence at the one month follow up as compared to the control group [*AOR* = 1.36 (*CI* = 1.16 to 1.61), *p* < .001]. Masaki et al. (2019) [25] reported that the overall continuous abstinence rate (CAR) results 76% (*CI* = 65 to 88) (12 weeks), 64% (*CI* = 51 to 76) (24 weeks), 58% (*CI* = 46 to 71) (52 weeks) were better than the results by the outpatient clinic (calculated through the national survey) and the varenicline (medication for smoking cessation) phase 3 trial in the United States and Japan. As secondary outcomes, Masaki et al. (2019) [25] reported decrease in social nicotine dependence (*M* = -6.7, *SD* = 5.2), tobacco craving (*M* = -0.6, *SD* = 1.5), and withdrawal symptoms (*M* = -6.4, *SD* = 5.8).

Calvaresi et al. (2019) [28] reported that 28.9% (78/270) participants completed their smoking cessation goal three months after the last cigarette. This result was 10% higher than the previous edition of smoking cessation program which was without the chatbot support. Carrasco-Hernandez et al. (2020) [27] reported that smoking abstinence (exhaled carbon monoxide (CO) and urine cotinine test) was 2.15 times [*CI* (1.13, 4.08), *p* = .02] higher in the intervention group than control group. However, none of the secondary clinical measures (health related quality of life, healthy lifestyle, and physical activity) showed a difference between groups. Conclusively, there was evidence indicating significant long term and short terms effects on smoking cessation through chatbot based interventions.

##### Substance Misuse

Out of the 15 studies, only 1 study aimed at reducing problematic substance use. Prochaska et al., (2021) [22] reported a significant increase in confidence to resist urges to use substances (mean score change +16.9, *SD* 21.4; *p*<.001), and a significant decrease in the following: substance use occasions (mean change -9.3, SD 14.1; *p*<.001), Alcohol Use Disorders Identification Test-Concise (mean change -1.3, SD 2.6; *p*<.001), 10-item Drug Abuse Screening Test (mean change -1.2, SD 2.0; *p*<.001), Patient Health Questionnaire-8 item (mean change 2.1, SD 5.2; *p*=.005), Generalized Anxiety Disorder-7 (mean change 2.3, SD 4.7; *p*=.001), and cravings scale (68.6% vs 47.1% moderate to extreme; *p*=.01).

##### Treatment or Medication Adherence

Out of 15 studies, 3 studies targeted medication or treatment adherence, but 2 studies reported the efficacy of AI chatbots in increasing treatment or medication adherence through timely and personalized reminders. Brar Prayaga et al. (2019) [31] reported that out of the total refill reminders (273,356), 17.40% (47,552) of the reminders resulted in actual refill requests. Furthermore, 54.81% (26,062/47,552) actually refilled within two hours of the reminder. Spanish-speakers had significantly lower refill request rates than English-speaking patients (10.69% vs. 18.83%) (*χ^2^* =1829.2; *p*<.001). Among these who engaged with the reminder, Spanish-speaking patients had higher refill request rates than English-speaking patients (57.31% vs. 49.22%, *χ^2^* =212.5, *p*<001). Younger patients (<45 years) had higher refill request rates than older (>75 years) patients (47.81% vs. 29.84%, *χ^2^* =1460.3, *p*<.001). Chaix et al. (2019) [20] reported that the average medication adherence rate improved by more than 20% in 4 weeks (*p*=0.4) through the prescription reminder feature. Conclusively, there was evidence indicating significant increase in medication adherence rate through the chatbot use, however, there were cultural differences observed in the chatbot usage.

#### Chatbot Intervention Characteristics

##### Behavioral change theories and expert consultation for intervention content

The chatbot intervention characteristics are summarized in Table 5. In more than half of the studies (*n*=9), the AI chatbots’ content, features, and interface were designed based on a theory. Each study critically selected theories based on the intervention goals and the target beneficiaries. The Cognitive Behavioral Therapy (CBT) was used in Tess, Lark Health Coach (HCAI), and Woebot to devise strategies that enhance self-efficacy and sustain behavior change. In Tess, CBT was clubbed with the theory on emotionally focused therapy and motivational interviewing to assist behavioral counselling of adolescent patients. Likewise, in Woebot, CBT was clubbed with motivational interviewing and dialectical behavior therapy to emotional support and personalized psychoeducation to resist substance misuse. The theory of motivational interviewing was also used to devise interview questions addressed by NAO (the social robot), and the motivation reinforcement messages by Bickmore et al.’s (2013) [24] Chat1. In HCAI, CBT was clubbed with the Diabetes Prevention Program’s (DPP) curriculum to develop content for conversations on weight loss.

**Table 5.**
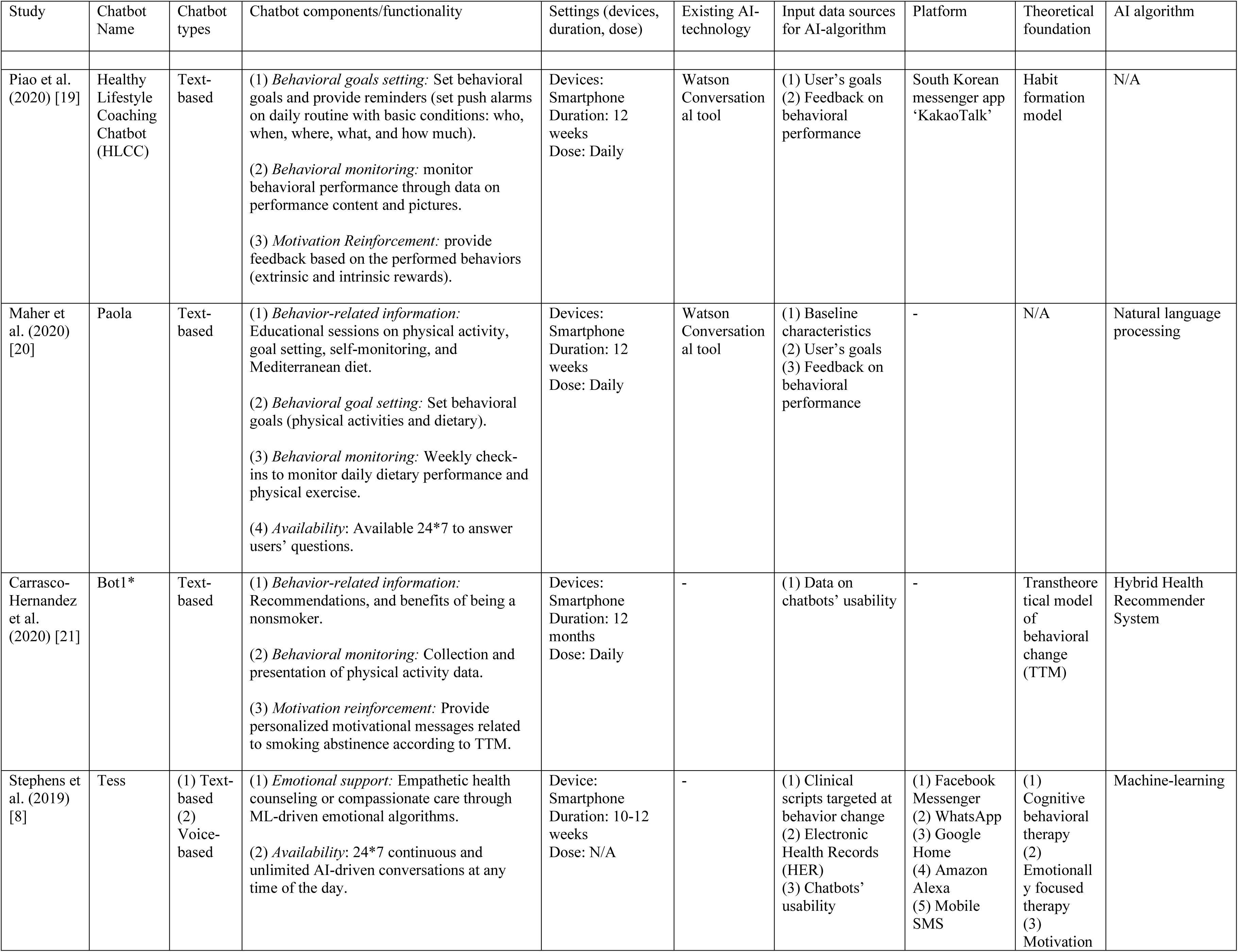
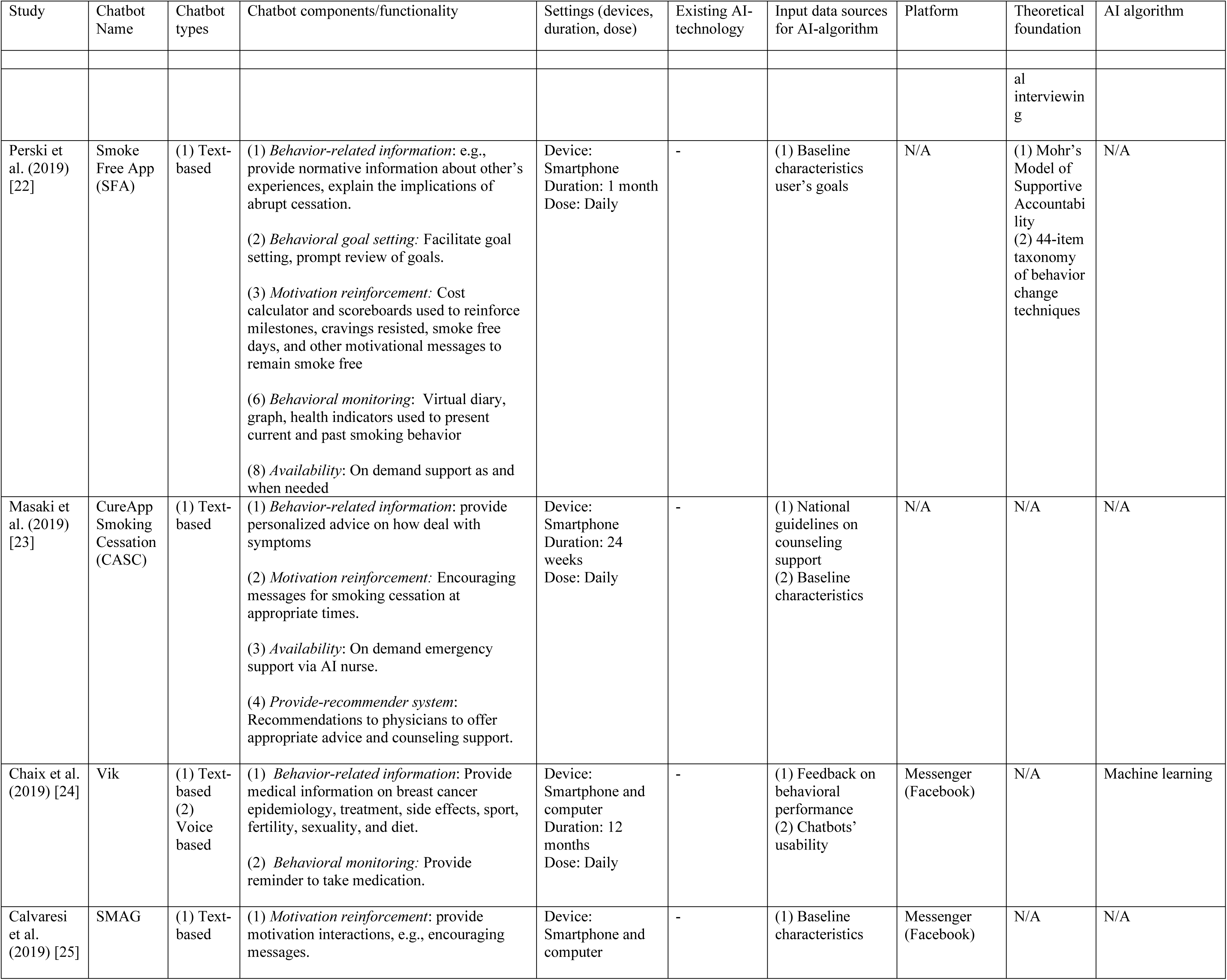
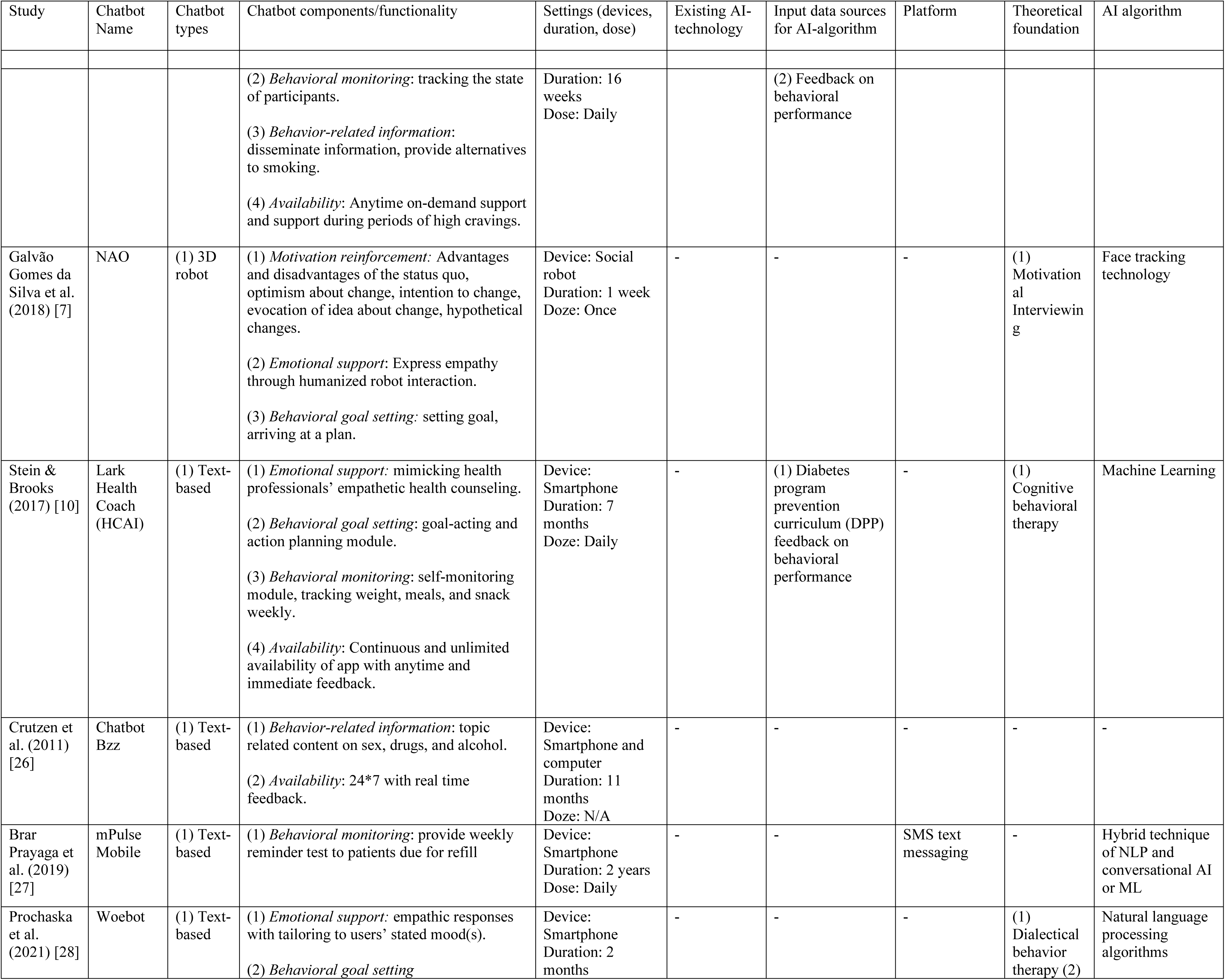
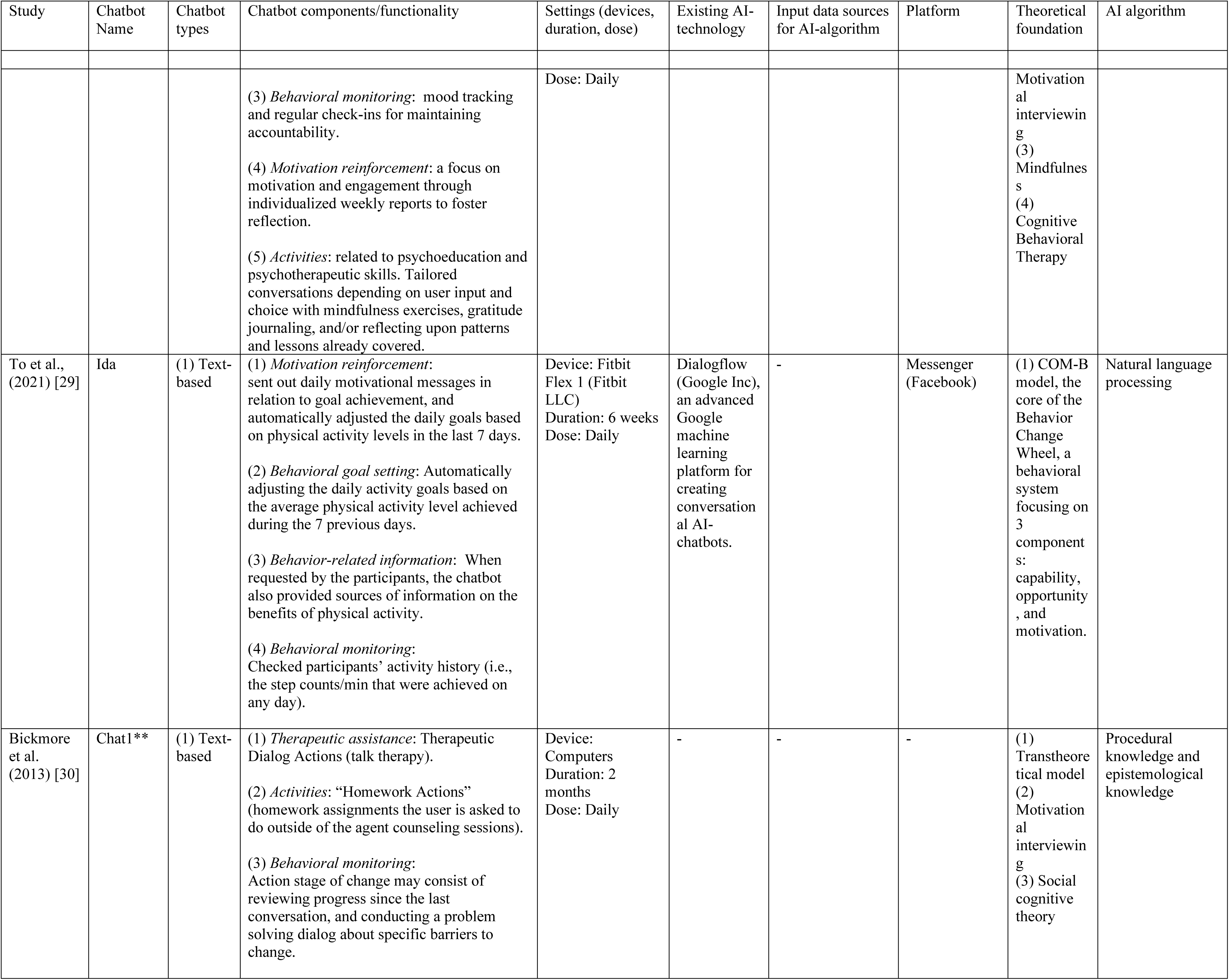
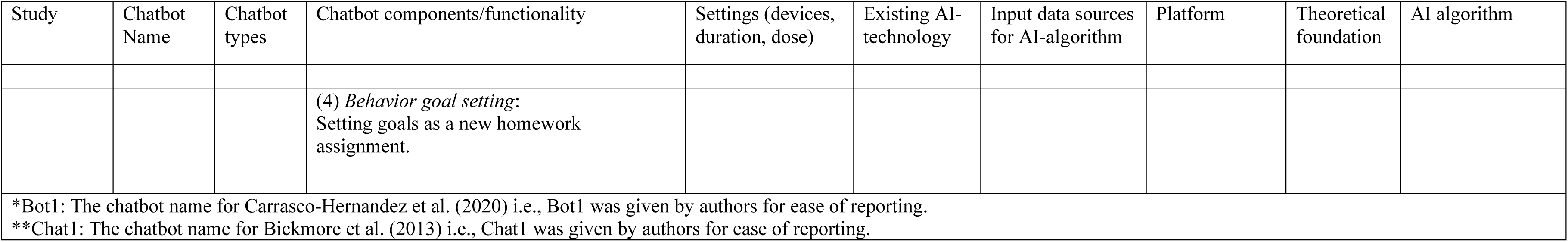
Chatbot features of reviewed studies

The habit formation model, which explains the relationship among Cues, behaviors, and rewards, was used to develop the reminder system in Healthy Lifestyle Coaching Chatbot (HLCC). The Mohr’s ‘Model of Supportive Accountability’, which states that inclusion of human support in digital interventions increases engagement, was used to mimic human support in Smoke Free App (SFA) to increase accountability and belongingness. Furthermore, SFA’s behavior change techniques were coded against a 44-item taxonomy of behavior change techniques in individual behavioral support for smoking cessation. The transtheoretical model (TTM) of behavioral change was used to decide message frequency by Carrasco-Hernandez et al.’s (2020) [27] AI-chatbot. Likewise, TTM was used in Bickmore et al.’s (2013) [24] Chat1 to design the behavioral monitoring process, which included reviewing progress, identifying barriers, and solving problems. COM-B model, the core of the Behavior Change Wheel, a behavioral system focusing on 3 components: capability, opportunity, and motivation, was used in To et al.’s (2021) [23] Ida to set goals, monitor, and reinforce behavior change through motivational messages. Social Cognitive Theory (SCT) was used in To et al.’s (2021) [23] Ida to facilitate the Therapeutic Dialog Actions (talk therapy), and homework sessions outside of the agent counseling sessions.

Apart from the use of theories, expert consultation and institutional assistance were adopted to develop AI-chatbots’ content. Mental health experts were consulted to develop and deliver customized messages through Tess, whereas, two national health promotion institutions in The Netherlands developed the content for Bzz. Conclusively, most studies either adopted a set of critically selected behavioral change theories or/and consulted with domain experts (individuals or/and institutions) to develop behavioral change strategies.

##### Chatbot functionalities

The AI-chatbots had multiple functionalities that contributed to efficacious outcomes. First, 8 studies targeted ‘behavioral goal setting’. These chatbots targeted healthy lifestyles (*n*=7, HLCC, Paola, SFA, NAO, HCAI, Ida, Chat1) and reduction of substance misuse (*n*=1, Woebot). The chatbots with goals related to healthy lifestules enabled users to set physical activity and dietary goals with push alarms to maintain daily routines and monitor weight, and. Second, 11 studies used ‘behavioral monitoring’. The chatbots that targeted healthy lifestyles (*n*=5, HLCC, Paola, HCAI, Ida, Chat1) enabled behavioral monitoring by consistently providing feedback through performance content and pictures, weekly check-ins and data-based performance feedback. The chatbots that targeted smoking cessation (*n*=3, Bot1, SFA, SMAG) offered data-driven feedback on health indicators through virtual diaries and graphs. The chatbots that targeted medication or treatment adherence (*n*=2, Vik, mPulse) offered timely reminders to take medications or refill medicines. The chatbot that targeted reduction in substance misuse performed mood tracking and regular check-ins for maintaining accountability (*n*=1, Woebot) Third, 8 studies offered ‘behavior-related information’. The chatbots that targeted healthy lifestyles (*n*=3) offered educational sessions benefits on physical activity (Ida) and healthy diet (Paola), information on sex, drugs, and alcohol (Bzz). The chatbots that targeted smoking cessation (*n*=4, Bot1, SFA, CureApp Smoking Cessation (CASC), SMAG) educated users on the benefits of being a nonsmoker, implications of abrupt cessation, and alternatives to smoking. The chatbots that targeted medication or treatment adherence (*n*=1, Vik) offered information on the health issue (breast cancer) for which the users were taking medication. Fourth, 8 studies reported ‘motivation reinforcement’. The chatbots that targeted healthy lifestyles (*n*=3) offered feedback on behaviors (HLCC, Ida), and reinforced optimism to change behaviors through planning and imagining change (NAO, Ida). The chatbots that targeted smoking cessation (*n*=4) reinforced motivation through personalized messages based on TTM (Bot1), scoreboards and trackers of milestones, (SFA) and motivational messages (CASC, SMAG). The chatbot that targeted reduction in substance misuse focused on motivation and engagement through individualized weekly reports to foster reflection (Woebot).

Fifth, 4 studies provided ‘emotional support’. Three studies targeted healthy lifestyles, and one targeted reduction in substance misuse. In healthy lifestyle interventions, Tess offered empathetic health counseling or compassionate care through ML-driven emotional algorithms; NAO, the social robot expressed empathy through humanized robot interaction; HCAI mimicked health professionals’ empathetic health counseling. In reduction in substance misuse, Woebot offered empathic responses by tailoring to users’ stated mood(s). Sixth, 1 study (CASC) delivered ‘provider-recommendation system’ services. CASC offered advice and counseling support to physicians. Seventh, 7 studies reported ‘24*7 availability’ of the AI-chatbot. The chatbots that targeted healthy lifestyles (*n*=4, Paola, Tess, HCAI, Bzz) offered on-demand support, unlimited conversations, and answers to infinite number of questions. The chatbots that targeted smoking cessation (*n*=3) offered on-demand emergency support via AI nurse (CASC), support during periods of high cravings (SMAG), and unlimited availability for conversations (SFA). Eighth, 2 studies promoted activities beyond conversation with chatbot. Chat1 offered homework assignments, whereas Woebot required mindfulness exercises, gratitude journaling, and/or reflecting upon patterns and lessons already covered. Conclusively, AI chatbots offered personalized, real-time feedback, and on-demand support to users continuously and indefinitely.

##### AI Techniques

Majority of the studies (*n*=10) deployed different AI-techniques to deliver personalized interventions: Natural Language Processing (NLP), Machine Learning (ML), Hybrid techniques (ML and NLP), Hybrid Health Recommender System (HHRS), Face-tracking technology, and procedural-knowledge and epistemological knowledge-based algoritm. ML- driven emotional algorithms were used in Tess and HCAI to provide empathetic counselling or compassionate care (emotion-based response). The AI-algorithm analyzed users’ messages (voice- or text-based) to identify and categorize user emotions. Thereafter, the chatbots provided both emotional and strategic support to the users. NLP and ML techniques were used in Paola, Vik, Ida, and Woebot to identify and categorize user intents and entities by analyzing unstructured messages. Bickmore et al.’s (2013) Chat1 used procedural and epistemological knowledge-based AI-algoritms that facilitated the therapeutic dialog actions (talk therapy).

Hybrid technique of NLP and conversational AI or ML were adopted by mPulse to ensure smooth, continuous, and uninterrupted conversation. HHRS was adopted by Carrasco-Hernandez et al.’s (2020) [27] AI-chatbot to personalize messages based on user demographics, content (interest of the user), and utility (ratings on each message by the user). Face-tracking technology was integrated into NAO (the social robot) to track participant’s face to humanize the interaction experience. The remaining chatbot studies (*n*=5) specified the use of artificial intelligence to personalize the chatbot interaction but did not elaborate on the AI-techniques adopted. Conclusively, most studies targeted ‘personalized services’ through different AI- techniques.

##### Logistics

The chatbots used multimodal channels of communication with the users. All chatbots except NAO (*n*=14) used text-based communication with the user, amongst which 2 chatbots (Tess and Vik) also used voice-based communication. NAO only used voice-based communication as it was deployed via social robot. The AI-chatbot-based interventions were implemented for different durations: 0-2 month (*n*=3), 2-5 months (*n*=7), 5-9 months (*n*=2), 9-12 months (*n*=2), and >12 months (*n*=1). Out of the 13 chatbots that reported frequency of engagement, all chatbots except NAO interacted with the users daily. NAO only interacted once because it was delivered in-person through a social robot.

The AI chatbots were either integrated into existing platforms or delivered as independent platforms. Vik, SMAG, Tess, and Ida were integrated to Facebook (FB) messenger. Tess was also available with WhatsApp, Amazon Alexa, Google home, and mobile SMS. HLCC was integrated with KakaoTalk, a popular messenger app in South Korea, and mPulse was integrated with mobile SMS. The remaining chatbots (*n*=8) were delivered independently. The chatbots were deployed using different devices. All chatbots except NAO, Ida, and Chat1 (*n*=11) were deployed through smartphones, among which 3 chatbots (Vik, SMAG, Bzz) were also deployed through computer. Chat1 was only deployed through computers. NAO was deployed through a social robot, and Ida was deployed through Fitbit Flex 1. Three chatbots (HLCC, Paola, and Ida) integrated an existing AI-driven conversational platform i.e., Watson conversation tool (HLCC, Paola) and Dialogflow, an advanced Google machine learning algorithm (Ida). Conclusively, AI-chatbots offer the opportunity to be deployed through accessible devices and platforms, indicating the potential for reaching remote and large populations.

##### Input Data for Personalized Services

To deliver personalized services using AI-chatbots, most chatbots/studies (*n*=9) required input data on user’s background, goals, behavioral performance, chatbots’ usability, and evidence-based content. User’s background information or baseline characteristics were collected by 4 AI-chatbots. Paola measured baseline level of physical activity and Mediterranean diet; SFA measured ‘time to first cigarette’ and ‘cigarettes per day’; CASC measured demographics, motivation levels for smoking cessation, number of cigarettes smoked per day, and years of smoking; SMAG measured demographics and type of smoking dependence; and Tess utilized electronic health records. Information on user’s goals i.e., who, when, where, what, and how, were collected by 3 chatbots. HLCC asked users (office workers) to set realistic stair climbing goals, Paola enabled users to set dietary goals and daily steps for every week based on the previous week’s outcomes, and SFA asked users to set the target quit date for smoking. Real-time feedback on chatbots’ usability was collected by 3 chatbots. Bot1 collected feedback on message content and message timing; Tess collected data on the usefulness of the message; and Vik collected data on the relevance of reminders.

Real-time feedback on behavioral performance of users was collected by 5 chatbots. HLCC collected performance content and pictures, Paola collected data on daily steps and dietary pattern, Vik collected data on medication adherence levels, SMAG monitored users’ smoking levels along with information on location, alone/accompanied, ongoing activity, and mood to create smoking profiles of users, and HCAI gathered data automatically through sensors on phones, integrated devices such as wearables, and self-reported information such as dietary consumption. Apart from user data, 3 studies used evidence-based content. Tess used clinical scripts targeted at behavior change, CASC used national guidelines on counseling support, and HCAI used content form the diabetes program prevention curriculum. Conclusively, most studies used diverse input datasets, indicating the need for collecting comprehensive and essential input data for delivering personalized services.

## Discussion

The results of this review demonstrate the potential of AI-chatbots to deliver efficacious, effective, and feasible health behavior interventions. However, the high risk of internal validity, lack of sufficient description of AI-techniques, and lack of generalizability of the selected studies suggest further research with robust methodologies to draw definitive conclusions. Regardless, the review identified practical or/and research implications of intervention strengths and limitations of the existing studies with potential future direction.

### Principle Findings

#### Primary Outcomes

This review found that AI-chatbots were efficacious in promoting healthy lifestyles (physical exercise and diet) (*n*=6), smoking cessation (*n*=4), treatment or medication adherence (*n*=2), and reduction in substance misuse (*n*=1). These findings are consistent with previous systematic reviews that reported the use of AI-chatbots for “improvement in physical activity levels” and “improvement in medication adherence” [2, 9]; “treatment adherence” [15]; “adherence to self-management practices” [1]; and “smoking cessation” and “reduction in substance abuse” [13].

#### Secondary Outcomes

Likewise, the review found that AI-chatbots demonstrated high feasibility, usability, acceptability, and engagement. Our findings on these secondary outcomes are consistent with findings from previous systematic reviews on AI-chatbots that reported “acceptability” [12, 15]; “engagement” [12,14,15]; “on-demand availability,” “accessibility,” and “high quality information” [12]; “satisfaction,” “helpful information,” “easy to use,” “informative,” and “trustworthy” [9].

### Implications of ‘intervention characteristics’

#### Theoretical foundation for behavior change

The fundamental characteristics of AI-chatbots played a critical role in determining efficacious outcomes. First, majority the studies (*n*=9), the use of critically selected behavioral change theories in the design and delivery of AI-chatbots contributed to efficacious outcomes. Our findings suggested that the integration of behavioral change theories such as CBT, TTM, motivational interviewing, emotionally focused therapy, habit formation model, and Mohr’s Model of Supportive Accountability resulted in the delivery of consistent motivational support to users through goal setting, monitoring or tracking behaviors, and reinforcement. These strategies not only contributed towards better primary and secondary outcomes, but also solved several challenges in the traditional face-to-face intervention models from users’ standpoint such as limited connectivity with the expert, lack of consistent motivation, and lack of access to diverse information overtime. Previous systematic reviews also reported that the use of CBT [2, 12], habit formation model, emotionally focused therapy, and motivational interviewing [2] for designing behavior change strategies via AI-chatbots contributed to better engagement, user motivation, and health behavior outcomes. More interdisciplinary collaboration between the behavioral health experts and computer scientists is needed to develop theorey-based AI-chatbots for behavioral change interventions.

#### Free flow conversation

Second, in all studies, the ‘free-flow conversations’ rather than ‘rule-based or constrained conversations’ with AI-chatbots enhanced user experience through personalization of services, delivery of diverse information, and the choice of user-initiated conversation. On the other hand, rule-based chatbots offer limited user-experience through constraints on the input data, finite set of conversations that are task-oriented and straightforward, and lack of user-initiated conversations. Thereby, significantly reducing the chatbots’ ability to provide a personalized touch or human-human interaction experience. This finding is consistent with previous systematic reviews that reported the need for a greater personalization in AI chatbots through feedback on user performance, accountability, encouragement, and a deep interest in user’s situation [14]. Likewise, Milne-Ives et al. (2020) [11] reported a need for greater interactivity or relational skills, empathetic conversations, and a sense of personal connection with user through compassionate responses. Therefore, it is critical for the AI-chatbots to establish appropriate rapport or relationship with the user through personalized and compassionate interactions for a sustained and engaging intervention. The AI experts need to establish pathways of comprehensive real-time data collection to produce accurate and personalized responses

#### Non-judgmental space

Third, in 3 studies, the humanistic yet non-humanistic construct of AI-chatbots provided a safe space for the users to discuss, share, and ask information on sensitive issues [7,20,21]. The ML-driven emotional algorithms offered the potential for perceiving and understanding human emotions [32], whereas, the ‘non-human interaction’ experience or the lack of interaction with a ‘real’ human made it easier for the user to self-disclose or confront sensitive information [33]. Thus, AI-chatbots demonstrate their potential in intervening with vulnerable populations, especially for stigmatized issues. For example: The period of adolescence is characterized with high social anxiety, therefore, adolescents perceive stigma in seeking services on sensitive issues such as mental health disorders, etc. In such scenarios, AI-chatbots offer sufficient privacy and anonymity for adolescents to express their thoughts and emotions freely. This finding is consistent with the previous systematic review that reported the use of anonymity in encouraging users to express their emotions freely [14].

#### Easy to be integrated into existing services

Fourth, most studies (*n*=8) reported that the AI-chatbots have a low threshold for integration into existing services yet a high reward. Most of the traditional behavioral interventions require in-person expert consultation or service delivery; however, this approach has several limitations from the implementor’s standpoint such as lack of consistent data collection, continuous monitoring, scalability, and sustainability of the intervention. AI-chatbots have a low threshold for integration into these traditional services because they do not put a strain on existing resources (experts, time, money, and effort). The chatbots can be freely deployed through daily-use platforms and accessed at any time by the users. The utilization of chatbot can integrate the behavior intervention into daily clinical setting and avoid additional burden on the existing healthcare providers. For example: Chatbots can independently offer low-intensity services such as information delivery to user. Furthermore, chatbots can offer provider-recommendation services, wherein, based on the analysis of real-time user data, the chatbots may offer ‘suggestions’ to the healthcare providers to offer more effective services [25]. Therefore, the public health professionals and healthcare providers can consider the integration of AI-chatbots into existing services or programs as a ‘support tool’ to the expert, rather than a ‘replacement’ [5].

#### Scalable

Fifth, most of the studies (*n*=10) had a large and diverse sample size, demonstrating the potential of scaling up chatbot-based interventions. Almost half of the studies had 200+ participants, with four studies consisting of a sample size ranging from 920 to 991217 participants approximately. Likewise, the selected studies not only included samples with diverse health and behavioral conditions (*n*=13) such as breast cancer, smoking, obesity, unhealthy eating patterns, lack of physical exercise, and medicate recipients, substance misuse, but also samples with no pre-existing conditions (*n*=2). This depicted the potential of AI-chatbots to reach a large and diverse population in different settings. This is because AI chatbots have the potential to be integrated into the extensively used existing platforms (text- SMS, Facebook messenger, WhatsApp) and deployed through commonly used devices (smartphones, computers, Alexa), making it highly feasible to access a large and diverse population. This finding is consistent with the previous systematic reviews that reported the integration of AI chatbots into diverse platforms such as Slack, messenger, WhatsApp, and Telegram [1,2,15], and the use of a large sample size such as >100 participants in 6/12 studies [5], and up to 454 participants [12]. Thus, public health professionals can deploy AI-chatbots for education, promotion of behavior change, and offer healthcare services to prevent health issues that impact a large population.

### Limitations of reviewed studies and future research directions

#### Nascent application of AI chatbots

Almost 75% (11/15) of the articles were published in the years 2019 and 2021, indicating that the use of AI-driven chatbot interventions for behavioral change is at a nascent stage. Most (*n*=9) of the studies adopted a pre-post study design with no control group with only 4 studies using RCT models, reinstating the immaturity in establishing causal connections between AI-based conversational agents and health behavior outcomes. This finding was aligned with many previous systematic reviews that reported 4/9 studies were RCTs (remaining were quasi experimental, feasibility, or pilot RCT studies) [5], 2/10 studies were RCTs (majority were quasi experimental) [15], and 2/17 studies were RCTs (majority were quasi experimental) [1]. The future studies need to adopt robust RCTs that can establish causal relationship between AI-chatbots and health outcomes.

#### Risk of internal validity

The outcome of this review needs to be interpreted with caution due to a moderate to high risk of internal validity within the selected studies. In the included studies, the risk of outcomes from unintended sources was high due to lack of information on measures to avoid the influence of other interventions and level of adherence to the intervention protocol, the risk of bias in the measurement of the outcomes was moderate to high due to lack of concealment of assigned intervention from evaluators and the lack of using validated and reliable outcome measures, and the risk of bias in analysis was moderate to high due to high dropout rates, lack of power calculation to estimate sample size, and lack of information on the use of intent-to-treat analysis. These findings are consistent with many previous systematic reviews that reported moderate risk of outcomes from unintended sources due to confounding in all quasi experimental studies [5], high risk of outcome measurement as evaluators were aware of the assigned intervention [5, 11] or non-validated instruments were used for outcome measurement [1, 12], and moderate risk of bias in analysis due to high attrition rate, lack of analysis methods for bias correction, lack of power analysis, and small sample sized at follow up [2, 5].

There was also inconsistency across studies in the measures of secondary outcomes (feasibility, usability, acceptability and engagement). This finding is consistent with most of the previous systematic reviews that reported heterogeneity in these secondary outcome measures across studies [1,2,5,6,9,12]. Firstly, this issue stems from the lack of common operational definition of secondary outcomes in context of chatbot-based interventions. Secondly, since the AI-chatbot intervention domain is relatively new, therefore, there are very less or no outcomes measures on feasibility, usability, acceptability and engagement with tested reliability and validity. Therefore, the researchers in selected studies had to develop their own measures for assessing outcomes. This led to inconsistency in the measures and their operational definitions across the studies. Future studies should shape the development of common ‘operational definitions’ for each of these outcomes to enable comparison and standardized reporting. Furthermore, the future studies should follow the NIH quality assessment criteria for controlled intervention studies [17] to assess their studies’ internal validity.

#### Lack of description of AI algorithm

In the current review, most of the studies (*n*=14) did not describe the characteristics and handling of the input data, along with other processes related to the AI-algorithm. This findings is consistent with the previous systematic literature review that reported “inconsistent usage of AI-software taxonomy” and “lack of depth of reported AI techniques and systems” [15]. In alignment with CONSORT-AI extension [18], the future studies need to elaborate on the following components related to the AI-algorithm: (1) the process of supplying input data to the AI algorithm. This includes the user interface that enables data collection, inclusion/exclusion criteria of input data, handling of unavailable data, and establishing the credibility of the data collected (for example: specifying source of input data); (2) the output by the AI algorithm, and the relevance of the AI output with the health-related goals; (3) elaboration on the AI functioning. The includes the type of personalization algorithm (ML, NLP, etc.), version of the AI algorithm, and the accuracy level of the algorithm; (4) errors or performance backlogs in the AI algorithm deployed. This would indicate the safety level of using AI-algorithms, especially with vulnerable populations; (5) the level and type of expertise required to integrate and successfully deploy the AI-algorithm; and (6) the skills required by the participants to use the AI chatbot. This would indicate the number of resources required and the feasibility level of using AI-algorithms.

#### Lack of generalizability

The selected studies were not representative of diverse geographies, cultures, and age-groups, which exerted a strong bias on the generalizability of the studies Out of the 13 studies that reported the geographical location, all (100%) were conducted in the developed world; majority (80%) of the studies were embedded in the western culture, apart from studies in Korea and Japan; and majority (>80%) of the studies were implemented with adults (≥18 years). These findings were consistent with the previous systematic literature reviews that reported all the chatbot intervention studies were conducted in developed countries [2,6,12,15], most studies were conducted with adults [2, 9], most studies did not focus on racial or ethnic minorities [2, 15].

To increase the generalizability in efficacy and feasibility of AI-chatbots, the future studies need to test their use in developing countries and with children and adolescents. The increased mobile connectivity and internet usage in developing countries [34] offer the potential of implementing AI-chatbot based health behavior interventions. The use of AI-chatbots can tackle the challenges faced by health systems in developing countries such as lack of experts, limited health infrastructure in rural areas, and poor health access [35]. Likewise, with the rise in use of smartphones and latest digital technologies among adolescents [36], AI-chatbots offer the opportunity to deliver engaging behavioral health interventions to them. The non-judgmental and non-stigmatic attributes of AI-chatbot based interventions offer a solution to the challenges faced by adolescents in seeking behavioral health services such as perceived and enacted stigma, and lack of motivation [5, 37].

#### Safety and ethics

In the current review, the evidence for patient safety was limited; however, the limited evidence stated that the chatbots were safe for behavioral and mental health interventions. Only one study i.e., Maher et al. (2020) [19] reported safety in terms of no adverse events. This findings was consistent with the previous systematic literature reviews that reported very few studies discussed participant safety or ethics in terms of adverse events or harms [1,2,5,9], and data security or privacy [2, 11]. The flexible, real-time, and large quantity of conversations with AI-chatbots increase the probability of error by the AI-algorithm. This can lead to unintended adverse outcomes especially in case of sensitive topics. Therefore, in context to the nascent use of AI technologies, future studies should assess and report the AI performance from an ethical and safety standpoint.

### Limitations of the current review

The systematic literature review has several limitations. First, a meta-analysis was not conducted for the reviewed studies. Due to heterogeneity in the research design, outcomes reported, and outcome measures, a meta-analysis was not perceived as feasible by the authors. Second, this review did not cover a comprehensive set of behavioral outcomes. The selected studies focused on only three behavioral outcomes: healthy lifestyle (physical activity, diet), smoking cessation, and treatment or medication adherence. However, this was also because the authors had adopted strict inclusion criteria for AI-chatbots, studies with rule-based chatbots were ruled out, restricting the number of behavioral outcomes covered. Third, articles from out of selected databases (e.g., Google scholar), unpublished work and conference articles, grey literature (e.g., government reports), and articles in other language were not included. Fourth, intervention studies that did not provide a clear description of AI-chatbots or are labeled AI-chatbots as key word were excluded.

## Conclusion

This review provides an evaluation of AI-chatbots as a medium of behavioral change interventions. Based on the outcomes of the selection studies (15), AI-chatbots were efficacious in promoting healthy lifestyles (physical activity, diet), smoking cessation, and treatment or medication adherence. The AI-chatbots also demonstrated feasibility, usability, acceptability, and engagement in diverse settings with diverse populations. The efficacious and effective outcomes of AI-driven chatbot interventions can be attributed to the fundamental characteristics of an AI chatbot: (1) personalized services, (2) non-judgmental safe space to converse, (3) easy to integrate into existing services, (4) engaging experience, and (5) scalable to a large and diverse population. However, the outcomes of this review need to be interpreted with caution because majority of the studies a moderate risk of internal validity. This is because the AI-chatbot intervention domain is at a nascent stage. The future studies need to test adopt robust RCTs, and provide detailed description of AI related processes. Overall, AI-chatbots have immense potential in integrating into existing behavioral change services due to (1) the ease of integration, (2) potential for affordability, accessibility, scalability, and sustainability, (3) delivery of services to vulnerable populations on sensitive issues in non-stigmatic and engaging manner, and (4) the potential for consistent data collection to support healthcare provider’s decisions.

## Data Availability

All data produced in the present work are contained in the manuscript

## Acknowledgements

Li X and Qiao S conceived the research topics and questions; TC and AA conducted literature searching and screening; AA, TC and Qiao S did the data extraction and analysis; AA and Qiao S developed the first draft; WD reviewed the paper and gave key feedback and edits. All the authors reviewed the manuscript.

Research reported in this publication was supported by the National Institutes of Allergy and Infectious Disease under Award #R01AI127203-5S1. The content is solely the responsibility of the authors and does not necessarily represent the official views of the National Institutes of Health.

The authors would also like to acknowledge the generous funding support from the University of South Carolina (USC) Big Data Health Science Center, a USC excellence initiative program [Grant No: BDHSC-2021-14] and [Grant No: BDHSC-2021-11].

## Abbreviations

HLCC: Healthy Lifestyle Coaching Chatbot
SFA: Smoke Free App
CASC: CureApp Smoking Cessation
HCAI: Lark Health Coach

---

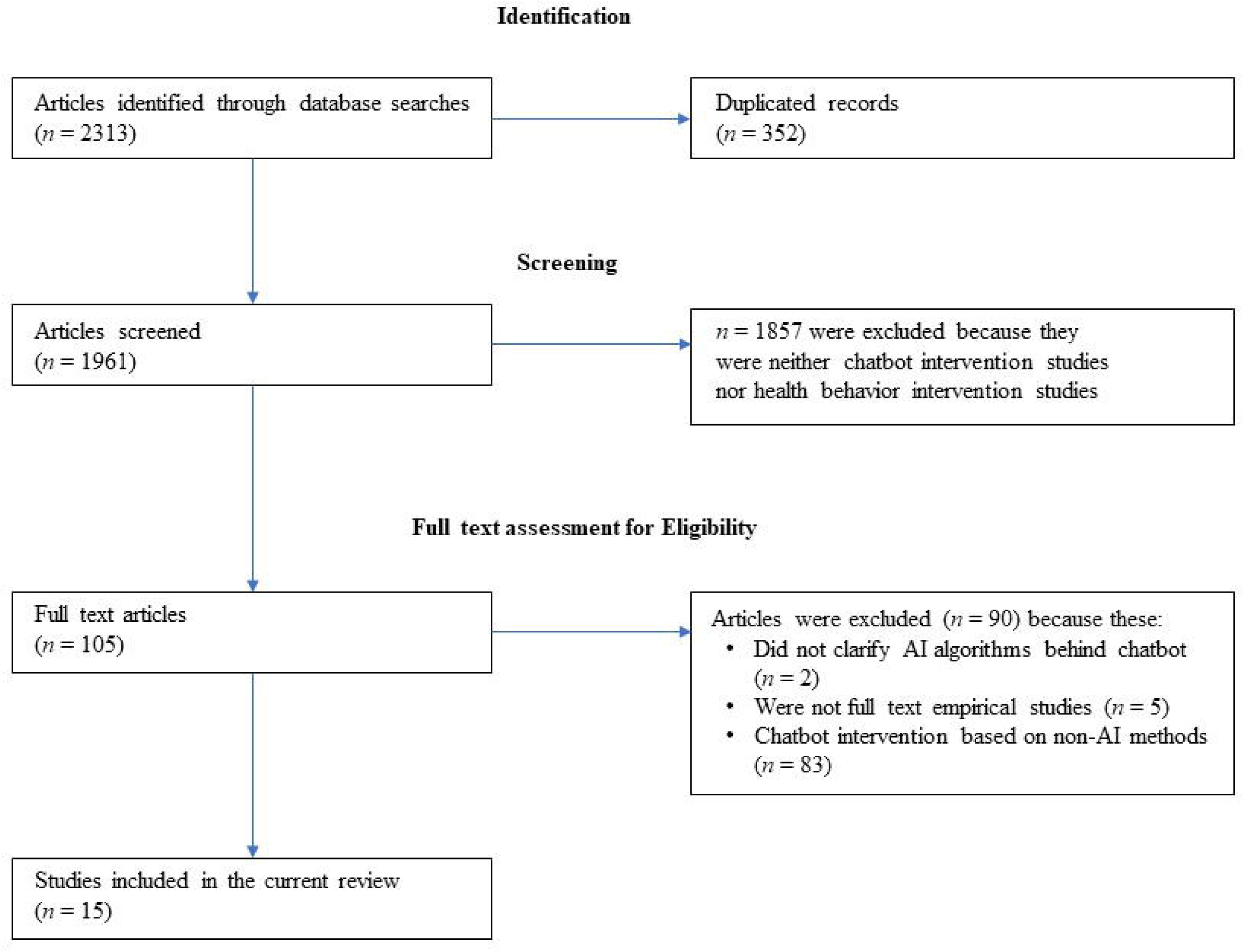

